# Are countries’ precautionary actions against COVID-19 effective? An assessment study of 175 countries worldwide

**DOI:** 10.1101/2020.07.16.20155515

**Authors:** Thamir M. Alshammari, Khalidah A. Alenzi, Fatemah A Alnofal, Ghada Fradees, Ali F. Altebainawi

## Abstract

**Background:** The Coronavirus Disease 2019 (COVID-19) pandemic has affected many countries negatively, particularly in terms of their health care and financial systems. Numerous countries have attempted to employ precautions to address this pandemic. This study was aimed at exploring and assessing the precautionary actions taken by 175 countries on six continents to prevent the spread of Severe Acute Respiratory Syndrome Coronavirus 2 (SARS-CoV-2).

**Methods:** An observational study was conducted based on data collected during the period from December 31, 2019, until the end of April 2020. Several data were extracted, including information related to the date of the first reported case of SARS-CoV-2, total confirmed cases, total active cases, and more. In addition, seven validated indicators were used to assess the countries’ preparedness and precautionary actions.

**Findings:** A total of 175 countries were included in the study. The total COVID-19 infection rate increased exponentially and rapidly in North America and Europe from March to April. The application of the precautions (indicators) varied between countries. School closures, quarantines and curfews were the most applied indicators among all countries. As for the relationship between the indicators and their effects on the infection rate, Italy and Spain were the top countries in Europe and adopted all indicators. Nevertheless, they faced high infection rates: 239,639 and 205,463 COVID-19 cases in Spain, and Italy, respectively.

**Interpretation:** The precautionary actions might have played a role in limiting the spread of COVID-19 in several countries. However, many countries did not benefit from applying these indicators.

**Funding:** No funding sources have been used for this work

## Introduction

During the last 20 years, there were several epidemics that were associated with viruses including SARS COV, H1N1 influenza and Middle East respiratory syndrome coronavirus (MERS-CoV) and all these epidemics imposes humanistic and economic burden on several countries.^1^ The first cases of coronavirus disease 2019 (COVID-19) was reported to the World Health Organization (WHO) in December 31, 2019.

The WHO initially believed COVID-19 would be limited to China. However, given the increased number of cases and countries that have been affected, it was considered a high-level epidemic.^1^ On March 11, the WHO declared COVID-19 a pandemic because it had spread to most countries, and millions of patients were affected by the disease worldwide.^2^

All data showed that, even in countries with good health systems, COVID-19 imposes a considerable burden not only on health care, but also at the country level. Countries such as Italy, Spain and the United States of America (USA), which are known to have good health care systems, have experienced a huge number of cases and deaths. In particular, the USA and United Kingdom (UK) account for a large percentage of deaths worldwide.^3^

These countries’ death rates ranged from 12.9% to 14.2%, which is considered high.^3,4^ These challenges make dealing with COVID-19 too difficult and could lead to huge burdens on the health care system.

The main difficult issues facing the health system in addressing COVID-19 include the fact that there is no available vaccine for Severe Acute Respiratory Syndrome Coronavirus 2 (SARS-CoV-2).^5^ However, several medications are currently used in an off-label manner for COVID-19. Among the main drugs used in this fashion are antimalarial medications (hydroxychloroquine and chloroquine), but their efficacy remains unproven.^6,7^

Therefore, the first method of addressing COVID-19 is preventing the spread of the virus. SARS-CoV-2 is highly transmissible among humans. This might be one of the reasons for the rapid spread of the virus and its becoming a pandemic.^5^

Several actions were taken at the source of the virus, Wuhan City, Hubei Province, China. These include isolation of any suspected or confirmed cases and their contacts, but the primary action taken was the restriction the mobility of the city’s residents.^2^ These actions are believed to delay the spread of SARS-CoV2.^8^ Moreover, the best indicators that might prevent or delay the spread of SARS-CoV-2 were taken by some countries as precautions before any cases occurred.^9^

Many countries took precautionary measures and actions aimed at reducing contact rates within the population and thereby reducing transmission of the virus. These included school closings, workplace closings and workforce reductions, public event cancelations, public transportation closures, public information campaigns, international travel restrictions, and quarantines and curfews intended to limit the spread of the virus.^10^ It is thought that containment indicators for COVID-19 may only slow its spread and that the virus is now entering a stage of unprecedented threat in terms of its global impact.^11^ However, these indicators are likely to be implemented to varying degrees depending on the countries in question and their strategies.^12^ The major challenge is maintaining the precautions and interventions until a vaccine becomes available.^13^ Therefore, this study is aimed at exploring the precautionary activities and patterns of 175 countries from six continents worldwide intended to address and prevent the spread of COVID-19.

## Methodology

### Study design

An observational epidemiological study was conducted based on data collected from all validated resources worldwide. The study included 175 countries from 6 continents worldwide (i.e., Asia, Africa, Europe, North America, Oceania and South America). The study was conducted during the period from December 31, 2019, until the end of April 2020. All countries’ information was searched, selected manually by the research team and divided by continents. After collection, the data were double-checked by the research team (each member checked the other members).

The country list was obtained utilizing the data from the WHO official page on the novel SARS□CoV□2.^14^ For each selected country, specific data related to SARS□CoV□2 and its indicators were collected. These included information related to the date of the first reported case, total confirmed cases, total active or suspected cases, total serious cases, total recovered cases, total deaths and deaths per million people starting from the beginning of the virus’ spread to the end of March 2020. In addition to total confirmed cases and deaths per million people, data such as number of confirmed cases, death rates, recovered cases and number of serious cases were also collected for April to compare the two months (March and April, 2020).

Validated indicators were used to assess the countries’ preparedness and precautionary actions. These indicators of government response included school closures (R1), workplace closures or workforce reductions (R2), public event cancelations (R3), public transport closures (R4), public information campaigns such as encouraging social distancing (R5), international travel restrictions (R6) and quarantines and curfews or movement reductions (R7).^15,16^

Several sources were used to extract the countries’ data, including the University of Oxford, Worldometer COVID-19 Data, KPMG International Cooperative, Health Ministries, UNESCO, the Government of the United Kingdom, and each country’s U.S. Embassy.

### Statistical Analysis

Descriptive statistics, including numbers and means, were used to compute government response indicators, infection cases, deaths, deaths per 1 million people and the percentage of increase in infections and deaths between March and April to obtain further insight into how countries’ actions affected the spread of COVID-19.

Data were exported to the Tableau software tool to support visual data analysis. The graphs created present the total number of actions in countries grouped by continent, total number of infections by continent, total deaths expressed as death rates in countries grouped by continent, total infection increase rate between March and April by continent, a comparison of the number of indicators taken with the number of deaths by continent and the differences in infections in March and April by continent. For data analysis, we used used IBM SPSS Statistics for Macintosh, Version 21.0.0.0 and Tableau Desktop, Version 2020.1.

## Results

A total of 175 countries were included in the study. Of these, 42 were located in Africa, 43 were in Asia, 5 were in Oceania, 47 were in Europe, 26 were in North America and 12 were in South America. All included countries applied at least one of the precautionary indicators. Infection rates were highest in Europe, followed by North America. Furthermore, Europe had the highest death rate at 6,524 per million people, whereas Oceania had the lowest at 8 per million people.

Worldwide, a maximum of 7 indicators were applied by governments. Figure A presents the total governmental indicators taken grouped by continent. Graph 1-A presents the total governmental indicators taken in 42 countries in Africa. Among these, 16 countries applied only 2 standards, 11 applied 3 standards, 2 applied 4 standards, 1 applied 5 standards, 8 applied 6 standards and 3 applied 7 standards. One country (Somalia) applied only one standard. The countries that applied all 7 indicators were Kenya, South Arica and Tanzania. All 43 countries closed schools, and only 20 countries closed workplaces and canceled public events. In total, 19 countries closed public transportation and used public information campaigns, and 11 countries applied international travel restrictions, as well as quarantines and curfews.

**Figure A.**
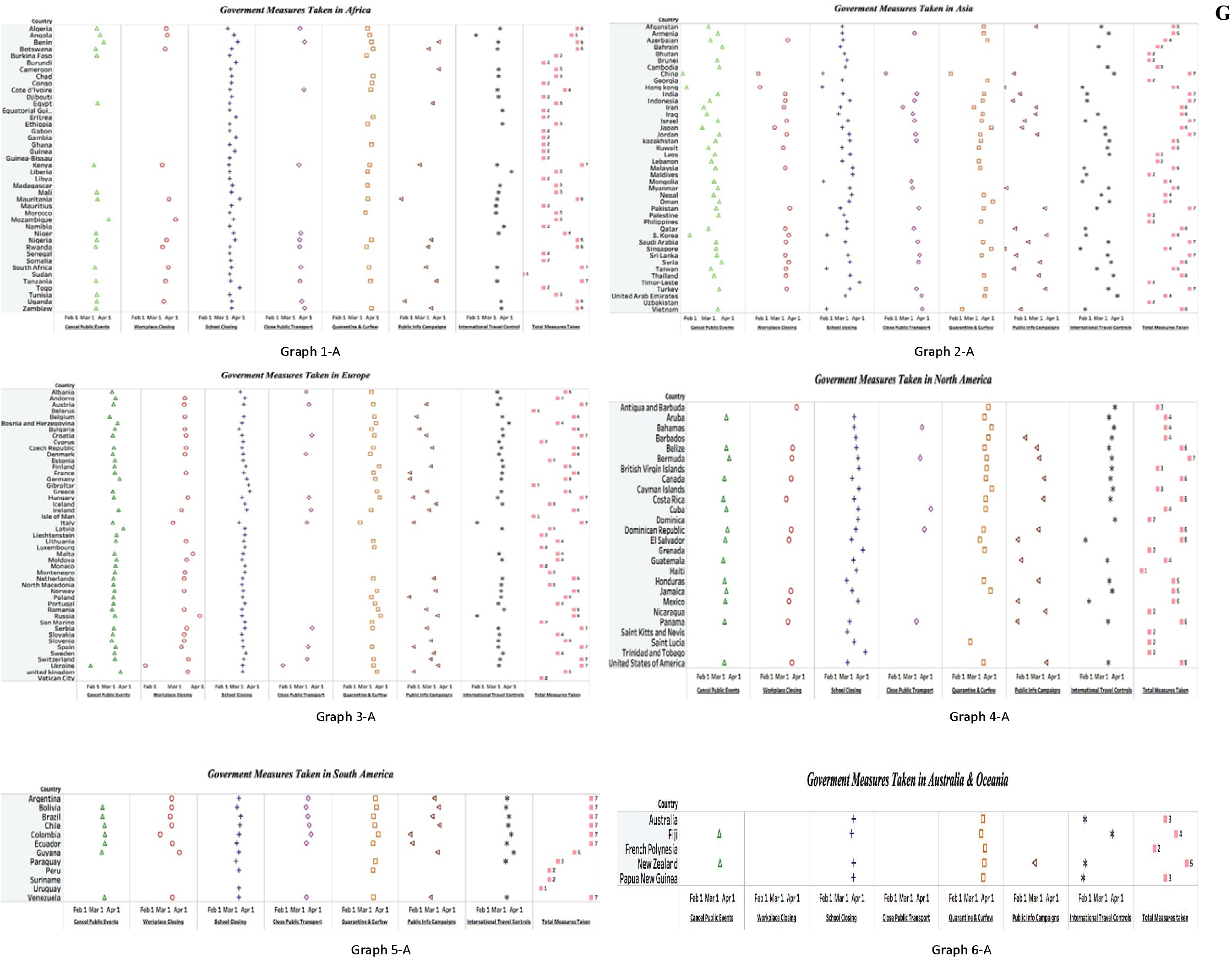
Government measures (indicators) that were taken by 175 countries.

Graph 2-A presents governmental indicators taken in Asia, where 43 countries applied precautionary indicators. Of these, 8 countries applied 2 indicators, 4 applied 3 indicators, 6 applied 4 indicators, 9 applied 5 indicators and 8 applied 6 and 7 indicators consecutively. Most countries closed schools, but only 23 countries closed workplaces. Furthermore, 36 countries canceled public events, and 19 countries closed their public transportation. A total of 21 countries used public information campaigns, 33 countries imposed international travel restrictions and 27 countries applied quarantines and curfews.

Graph 3-A shows the total governmental indicators taken in 5 countries in Oceania. Among these 5 countries, 1 country applied 2 indicators, 2 applied 3 indicators, 1 applied 4 indicators and 1 applied 5 indicators. No countries applied 6 or 7 standards. Most countries closed schools, imposed international travel restrictions and established quarantines and curfews. Only New Zealand and Fiji canceled public events.

Graph 4-A presents the total governmental indicators taken within 47 countries in Europe. Among these, 3 countries applied 1 indicator, 6 applied 2 indicators, 5 applied 3 indicators, 8 applied 4 indicators, 7 applied 5 indicators, 10 applied 6 indicators and 8 applied 7 indicators. Austria, Croatia, Hungary, Italy, Serbia, Spain, Switzerland and Ukraine applied all 7 indicators. Most countries closed schools and canceled public events, and almost half closed workplaces and used public information campaigns. Only 11 countries closed public transportation, whereas 36 imposed international travel restrictions and 30 established quarantines and curfews.

These governmental measures or indicators were a bit different in North America. Among 26 countries, 1 applied 1 indicator, 6 applied 2 indicators, 3 applied 3 indicators, 5 applied 4 indicators, 3 applied 5 indicators, 7 applied 6 indicators and 1 applied 7 indicators. Only Bermuda applied all 7. Most countries (24) closed schools, but only 13 closed workplaces and canceled public events. Only 5 countries closed public transportation, whereas 13 countries used public information campaigns; 18 imposed international travel restrictions and established quarantines and curfews. For further details, see Graph 5-A.

Graph 6-A presents the total governmental indicators taken in 12 countries in South America. Among these, 1 country applied 1 indicator, 2 applied 2 indicators, 1 applied 3 indicators, 1 applied 5 indicators, 1 applied 6 indicators and 7 applied 7 indicators. Most countries (11) closed schools, and the majority (9) closed workplaces, imposed international travel restrictions and established quarantines and curfews. Seven countries closed public transportation and used public information campaigns, and 7 canceled public events.

Another important factor was the total death rate in the studied countries. Figure B shows a plot chart of total deaths per million people versus total infections on 6 continents. Graph 1-B shows a plot chart of total deaths per million people versus total infections in Africa during the period January 2020–April 2020. Algeria had the highest death rate at 10 per million people and 4,006 infections, followed by Benin, which had 8 deaths per million people and a total of 64 infections. Morocco had a death rate of 5 per million people and a total of 4,423 infections.

**Figure B.**
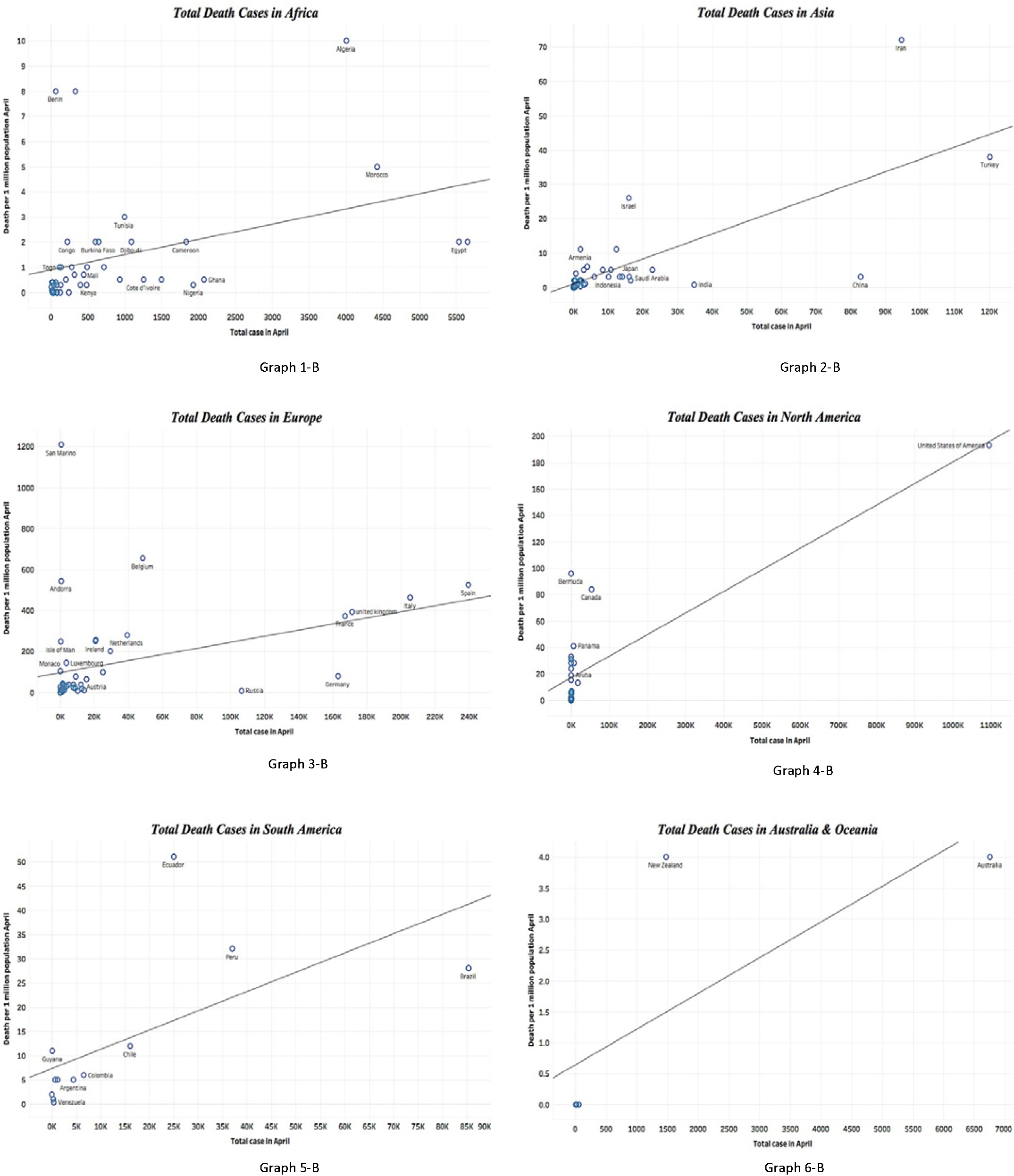
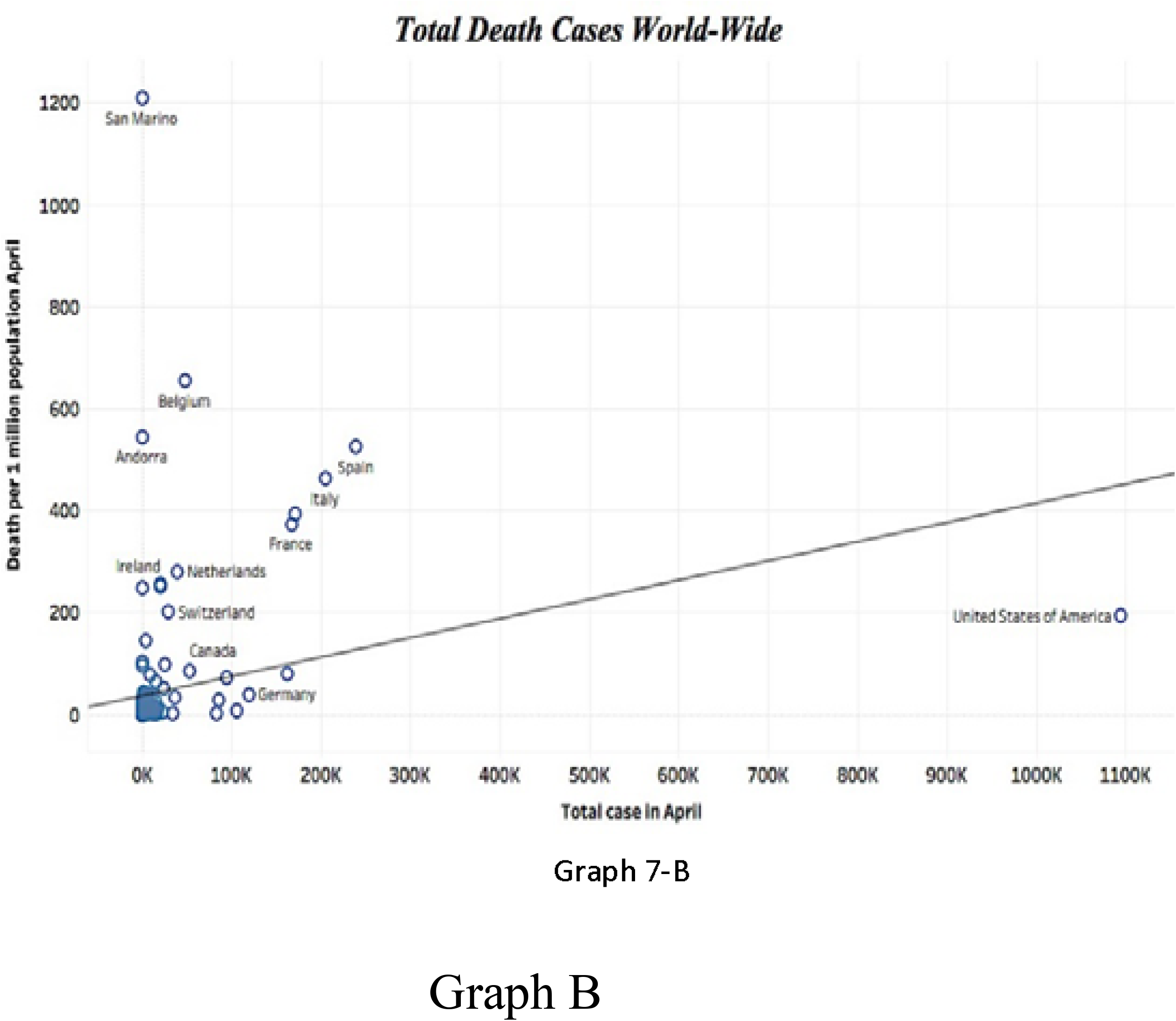
Number of total COVID-19 death cases in the 6 continents and worldwide.

In Asia, Iran had the highest death rate at 72 per million people and 94,640 infections. Turkey’s death rate was 38 per million people, and the country had 120,000 infections. Israel had a death rate of 26 per million people and 15,946 infections. For further details, see Graph 2-B.

Graph 3-B shows the death rate in Europe, where San Marino had the highest death rate at 1,208 deaths per million people and 569 infections. Belgium had a death rate of 655 per million people and 48,519 infections. Spain had the highest total infections and a death rate of 525 per million people.

The USA had the highest death and infection rates in North America with 193 per million people and 1,095,023 infections. Bermuda was second with a death rate of 96 per million people and 114 infections, graph 4-B. In neighboring South America, death rates ranged from 0.4 to 51. Ecuador’s death rate was the highest at 51 deaths per million people and 24,934 infections. However, Brazil had the highest infection rate with 85,380 cases and a death rate of 28 per million people, graph 5-B. In Oceania, Australia and New Zealand had death rates of 4 per million people, with 6754 cases in Australia and 1476 in New Zealand. For further information, graph 6-B.

As shown in graph 7-B, worldwide, the death rate varied from 0.06 to 1200 deaths per million people. Bhutan in Asia had the lowest death rate of 0 per million people and only 7 infections. In Europe, Vatican City had the lowest death rate at 0 per million people and only 11 infections.

Figure C compares countries and their total indicators taken using total infections between March and April 2020. In Africa, South Africa had the highest number of infections during March (1,353), followed by Algeria (716). These countries applied 7 and 6 indicators, respectively. However, by April, Egypt and Morocco had joined South Africa and Algeria due to a considerable increase in their number of infections. With 5,537 and 4,423 infections, respectively, and 3 indicators taken, both countries had a relatively similar number of infections compared with South Africa and Algeria (5,647 and 4,006, respectively). For further details, see Graph 1-C.

**Figure C.**
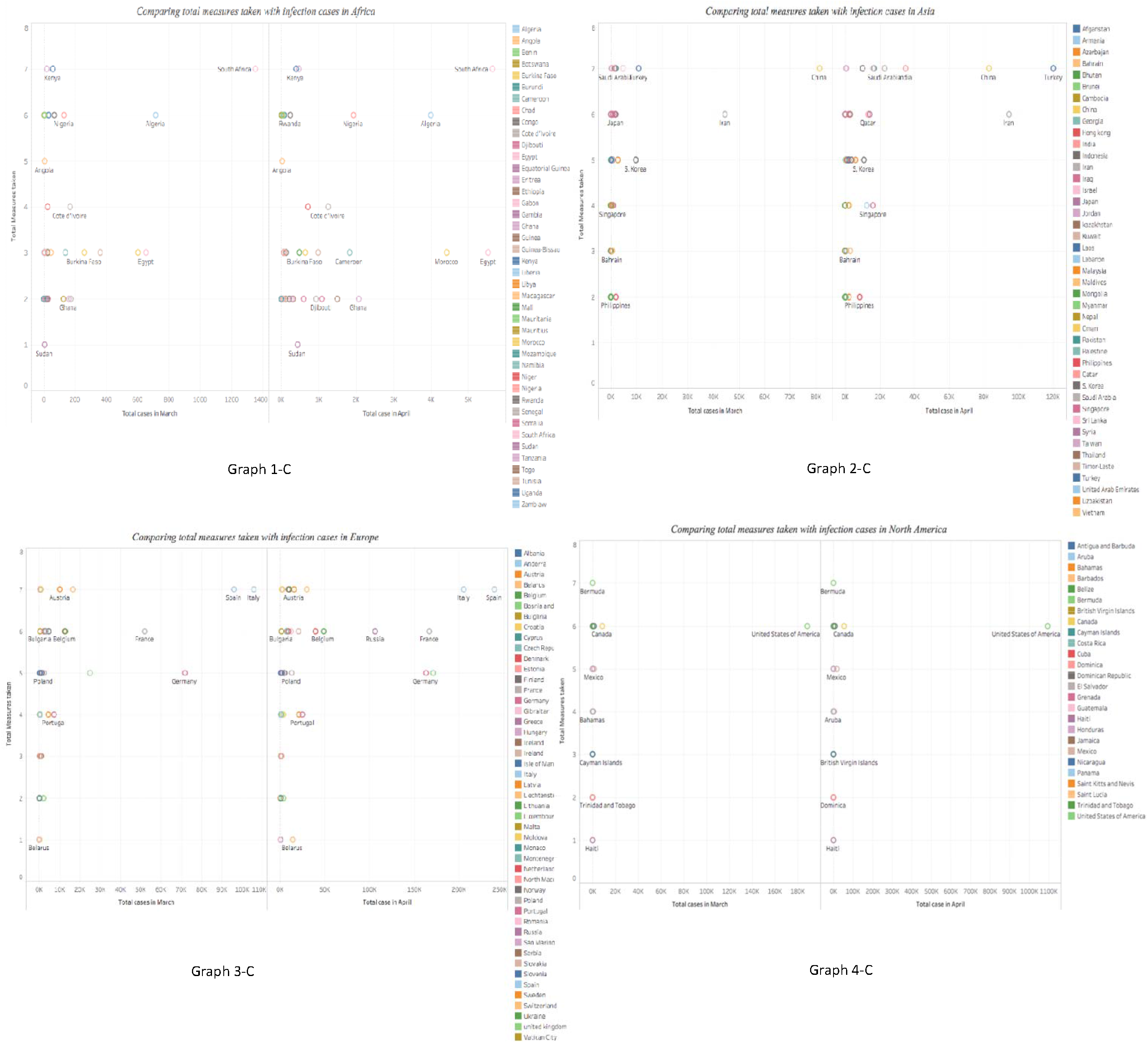
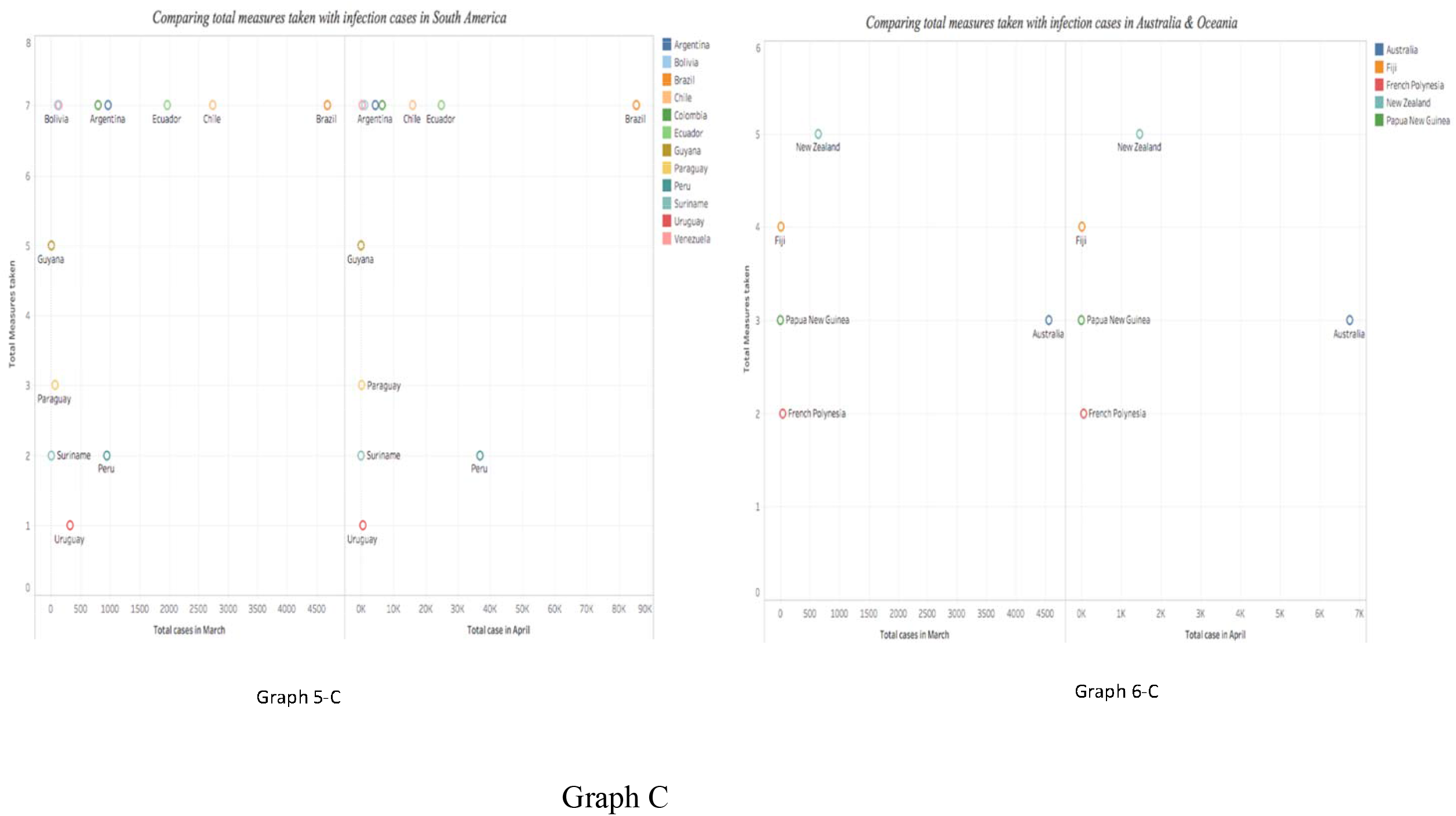
Comparison of number of death rate and total measures.

In Asia, China and Iran had the highest number of infections in March (81,518 and 44,605, respectively) and 7 and 6 indicators taken, respectively. However, by April, cases in Turkey (120,204), which applied 7 indicators, outnumbered those in China. For further details, see graph 2-C.

Graph 3-C presents infections rates in Europe, where most counties showed an increase during both months. Spain and Italy adopted all 7 indicators but faced the highest infection rates. In North America, as Graph 4-C shows, the USA, which applied 6 indicators, had the highest infection rate during March and April.

As Graph 5-C shows, in South America, Brazil applied all 7 indicators and had the highest infection rate in March and April. However, Uruguay applied only 1 indicator and had the lowest number of infections on the continent.

In Australia and Oceania, as Graph 6-C shows, a steady increase of infection cases occurred continent-wide. However, Australia had the highest infection rate and applied only 3 indicators. For further details, see Figure C.

In a comparison of the COVID-19 death rates among continents, Europe and North America had the highest, whereas Oceania and Africa had the lowest (see Graph 2-D). In addition, the total COVID-19 infection rate increased exponentially and rapidly in North America and Europe from March to April (see Graph 1-D).

Figure E presents a stacked bar chart views that shows that the number of infections in Asia increased during the period March-April 2020. As of March 30, the total number of infections in Asia was 175,130. By the end of April, this number had reached 510,711 reported cases (see Graph 2-E). However, compared to Asia, Africa’s rate of increase was greater. In total, 5,412 cases occurred in Africa, but the total number of infections rose to 37,631 by the end of April 2020 (see Graph 1-E). Countries in Oceania were not affected in both months, and their rate of increase was low, as the total number of cases was 5,250 at end of March and 8,314 at the end of April (see Graph 6-E).

**Figure D.**
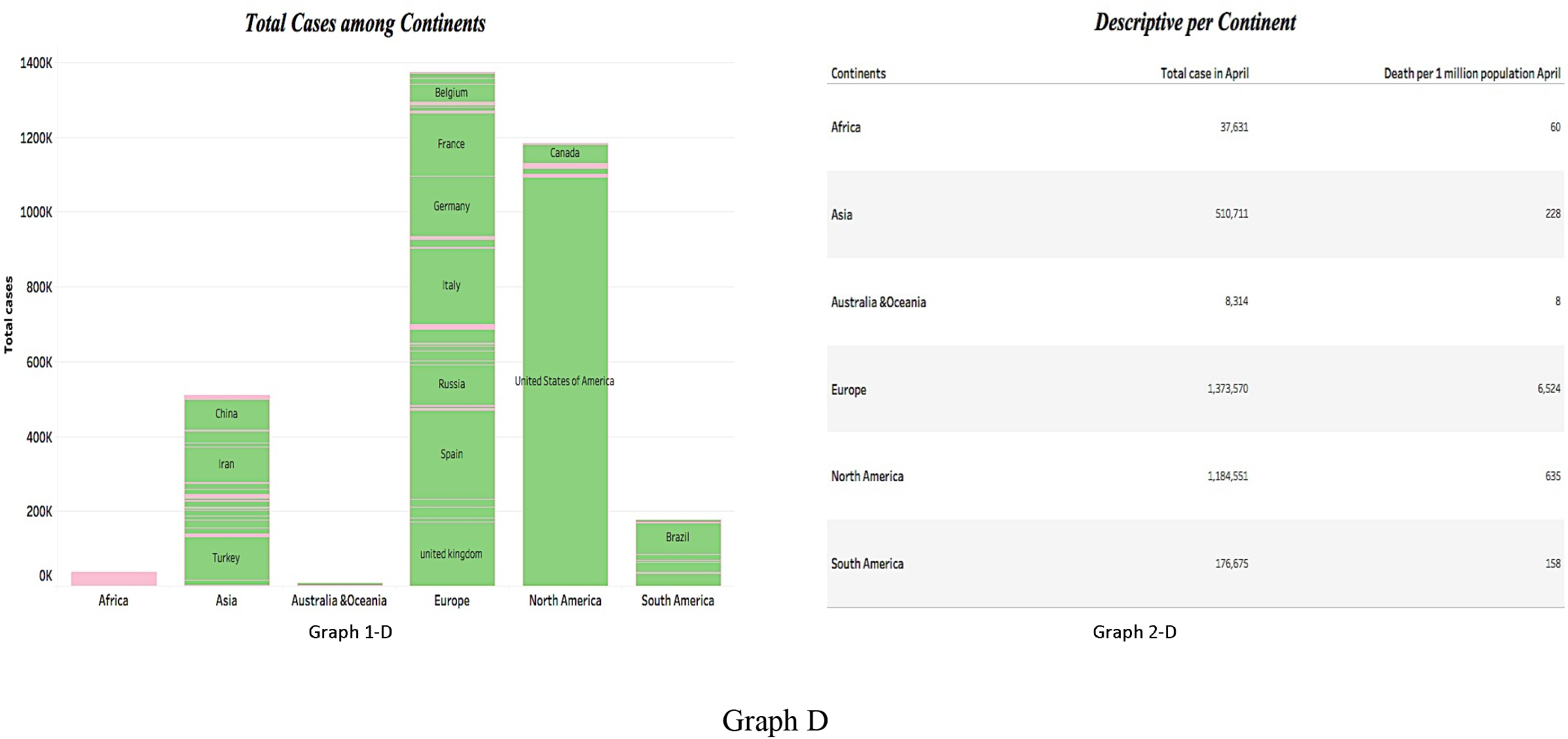
Total number of COVID-19 cases and death rate per million among the six continents.

**Figure E.**
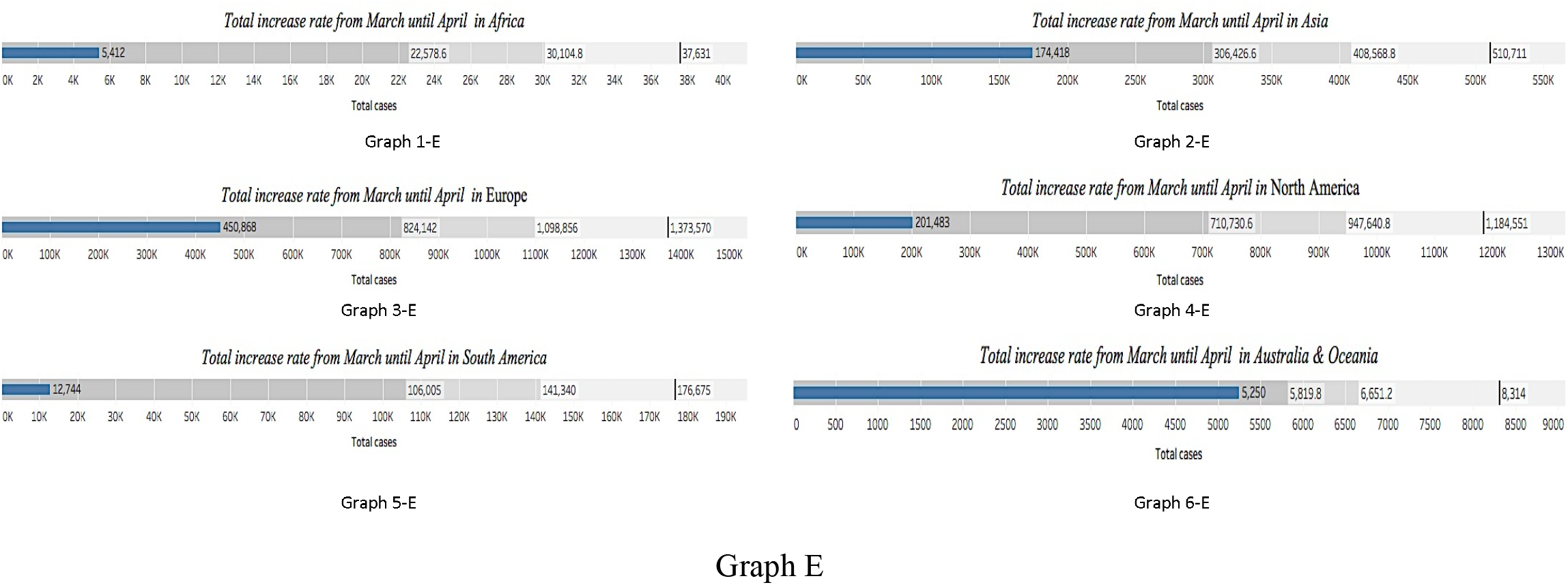
Increase rate of COVID-19 cases from March to April, 2020 among the six continents.

**Figure F.**
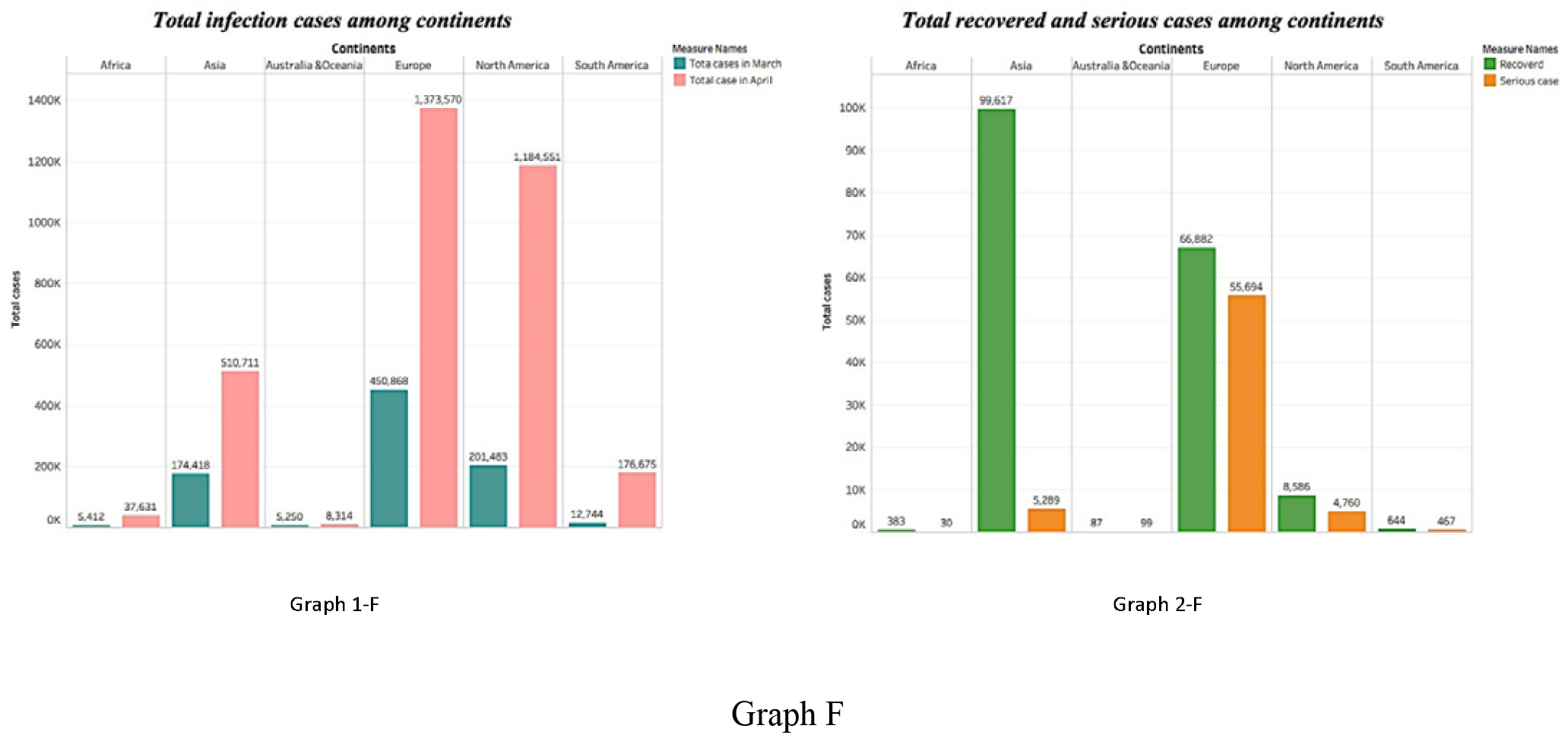
Total number of COVID-19 active, serious and recovered cases among the six continents.

In March and April 2020, the number of cases in Europe and North America increased by 6 and 3 times, respectively (see Graphs 3-E and 4-E). As of March 30, the total number of infections in North America was 201,483, but by the end of April 2020, it had reached 1,184,551. Europe had 450,868 cases on March 30 and 1,373,570 by the end of April.

The highest rate of infection increase was observed in South America, which increased by 13 times. The total number of infections was 12,744 at end of March, but this increased to 176,675 by the end of April 2020 (see Graph 1-F). The recovered cases pattern was similar for all continents except Europe, as the number of serious cases was 55,694 there but far lower elsewhere (see Graph 2-F).

## Discussion

On December 31, 2019, Chinese authorities alerted the WHO to an outbreak of SARS□CoV□2 in Wuhan City, Hubei Province, China. On March 11, 2020, the WHO announced that COVID-19 had become a pandemic. The rapid spread of COVID-19 worldwide has led to the creation and adoption of a wide range of responses in various countries to address COVID-19 outbreaks and reduce severe global socioeconomic disruption.

Recommended preventive indicators included hand washing, covering one’s mouth, maintaining distance between people and self-isolation for those who suspect they are infected. However, the governments differed widely in which measures they adopted and how quickly they implemented changes such as school closures, workplace closures and workforce reductions, public event cancelations, public transportation closures, public information campaigns, international travel restrictions (travel to or from China or all travel) and curfews, resulting in the uneven spread of the virus.

In general, government responses have become more stringent over the course of the COVID-19 outbreak. However, responses also vary among countries. This difference may be due to fear of economic repercussions, weak or strong resources, and wars.

This study examined the actions taken by 175 countries on 6 continents worldwide. We aimed to cover most regions worldwide and investigate the patterns among these countries. Thus, there were no specific exclusion criteria for countries, but inclusion depended on the availability of and access to data. Although COVID-19 originated from China, the highest death rate was in Europe, not Asia.^17^ This could have occurred for many reasons. The number of COVID-19 cases was high during the study period, and Europe’s elderly population is quite large; furthermore, most COVID-19 deaths occurred among elderly people due to weak immune systems.^18,19^ In addition, a lack of some important medical equipment such as test kits, personal protective equipment, ventilators and available beds in intensive care units in some countries, especially those with very high numbers of COVID-19 cases, might be another reason for this high death rate.^20^ In contrast, Oceania had the lowest rate at 8 per million people. This might be due to the distance between the continent’s islands, its low number of population, and the early precautionary actions on the part of some of countries in Oceania, which limited the number of COVID-19 cases in these countries. New Zealand is a successful example of controlling the COVID-19 pandemic. The number of tests conducted indicates that testing was performed more than once,^21^ and seven indicators or indicators were used to assess the country’s activities and preparedness to fight SARS-CoV-2. Only 15% of countries applied all 7 indicators, whereas the largest proportion applied only 2 indicators (22%). However, this pattern is different for Africa, South America and Oceania. Fewer indicators were applied by a couple of countries in Oceania. Thus, this continent had the lowest rate of COVID-19 pandemic infection and the fewest governmental indicators. This does not indicate that these indicators had no effect, however. These results could have occurred for several reasons: 1) applying some indicators such as quarantines and school closures early, 2) closing the country by preventing international travel to and from some countries and 3) Oceania’s location, which, unlike Europe’s makes travel between countries difficult because of their sea borders.^21^ These reasons could also be why the spread SARS-CoV-2 was lowest in Oceania. When comparing cases at end of March to those at the end of April, we found that the rate of increase was only 36% compared to 74% worldwide. Similarly, the death rate was increased by about the same percentage.

However, cases differed across other continents, where SARS-CoV-2 spread more when governments applied fewer indicators. This is clearly shown in data for continents such as South America, Africa and North America, where the rate of spread was higher than the worldwide rate. Asia and Europe were comparable, and both continents’ rate of spread was lower than the worldwide rate, possibly due to their extensive efforts in applying most indicators to limit the spread of SARS-CoV-2.

The spread of SARS CoV-2 among the studied countries might have been affected by the applied indicators; this was seen in various countries. In Oceania, the more indicators governments applied, the fewer COVID-19 cases they reported. For example, in New Zealand and Fiji, this trend continued for both months (i.e., March and April). Several other continents’ countries exhibited the same pattern, such as Kenya, Saudi Arabia, Japan, Austria, Switzerland, Bermuda, Canada, Bolivia and Argentina. However, this was not always the case because some countries applied more indicators but still faced high numbers of infections, as in China, South Africa, Spain, Italy and the USA. This might have occurred because these indicators were not followed well, were applied late after many cases had already occurred, or were not well recognized of being of considerable importance especially during the early period of the pandemic, such as in China.^22,23^

When comparing the indicators or measures used by continents with the mortality rate during the end of the first month, in Africa, most countries that applied more indicators, such as Kenya, Mauritania, Angola and Cote d’Ivoire, had lower mortality rates. However, results were different in Algeria, which had a high mortality rate despite applying 6 of the 7 indicators. The only indicator Algeria did not apply was the public information campaign, which might indicate that the educational materials and information about SARS-CoV-2 played a major role in this pandemic. Similarly, in Europe, Switzerland and Germany had low fatality rate, and both countries applied high number of these indicators.^24^ However, countries such Spain and Italy also applied high numbers of indicators but still had high mortality rates. This might be due to late application or lack seriousness in applying these indicators by these countries.^16,25^ In addition, as mentioned earlier, demographics and aging might have played roles in these countries.^20^

When the mortality rate was compared among countries worldwide, most countries with higher fatality rates were in Europe. The country with the highest rate was San Marino, followed by Belgium, Andorra, Spain, Italy, the UK, France and Sweden. This might be because of the massive number of COVID-19 cases that occurred earlier in Europe, as hospitals might not have been ready to deal with this high number of cases.^20^ Another explanation could be the aging population in these countries, especially considering that such individuals are part of the most affected age group.^26^

There were more serious COVID-19 cases in Europe compared to comparable continents. This sharp increase in cases occurred mainly in Italy and Spain, possibly due to the unpreparedness of these countries’ health care systems, especially when faced with a huge number of cases. In addition, these countries’ populations contain more elderly people, and most of the affected people were elderly; this could be another justification for such a high number of serious cases.^20,26^

When comparing total SARS-CoV-2 infection rates among continents for the months of March and April, most countries continued to increase at the same rate for both months regardless of the number of governmental indicators taken. However, in Asia and Europe, Turkey and Spain had an increase of percentage higher than the average infection rate, overtaking China and Italy, which were considered the most infected countries in Asia and Europe.

Several actions were taken at the source of the virus, Wuhan City, Hubei Province, China, including improved rates of diagnostic testing, immediate isolation of any suspected or confirmed cases and contacts and the restriction the mobility of the city’s residents.^27^ These actions are believed to delay the spread of SARS-CoV2.^28^

Some countries such as Singapore, Hong Kong and Taiwan relied more on early precautions because of their belief that prevention was the only option without an available vaccine or drugs. Singapore was among the first countries to apply a travel ban from China in late January. In addition, daily testing of up to 2000 people was conducted to check for SARS-CoV-2, and employees were asked to work from home due to mandatory quarantines in the country. Similar actions were taken by Hong Kong, and most people believed in these actions because the country was affected by SARS-CoV during the 2003-2004 pandemic.^29^

Some indicators were reduced in countries in late April due to a number of reasons. The outbreak of the pandemic affected educational systems worldwide, leading to the near-total closures of schools and universities and causing a destabilizing threat to the global economy. In addition, the increased global use of equipment to combat virus outbreaks caused food and supply shortages. The SARS-CoV-2 outbreak has had various impacts worldwide, especially in terms of disrupting factories and logistics operations.^30^

This study has several advantages. To our knowledge, it is the first study to evaluate these 7 indicators as precautionary actions on the part of 175 countries worldwide. In addition, our study involved important indicators that cover the most actions taken by countries. It also included data such as the number of infections, number of serious cases, number of recovered cases, number of deaths, death rate per million people and number of indicators applied by each country. The study’s limitations include the fact that we lacked time series information (i.e., the number of infections or deaths for multiple time points) because some data were unavailable or difficult to acquire.

## Conclusion

Most studied countries exhibited the same pattern in terms of increasing numbers of COVID-19 cases. This study showed that the precautionary indicators taken by various countries might have played a role in limiting the spread of COVID-19. However, some countries did not benefit from applying these indicators.

## Data Availability

The data are available with the corresponding author he will make them available whenever needed

